# Scoping Review: Differences between the risk factors of indigenous and non-indigenous latin american and caribbean children younger than 5 years old

**DOI:** 10.1101/2023.05.08.23289688

**Authors:** Daniel David Rodriguez Romero, Ana María Rojas Gómez, Jesús David Cuadrado Guzmán

## Abstract

**Objective:** To map the existing literature on the difference in risk factors for undernutrition between indigenous and non-indigenous children younger than 5 years old of Latin America and caribbean.

**Introduction:** Compared to non-indigenous children, Indigenous children have a higher risk for undernutrition, however, there are not systematic or scoping reviews identifying this difference through the risk factors.

**Inclusion criteria:** Every paper designed to assess directly or indirectly associations between undernutrition and risk factors, also, it must show results where indigenous people had not been aggregated to other races. We excluded every paper that didn’t have a covariate different to ethnicity.

**Methods:** This scoping review will use the JBI methodology to search several databases, trial registries, and gray literature for both published and unpublished studies related to malnutrition. Two or more independent reviewers will screen titles and abstracts for inclusion criteria, assess full texts, and extract data using a data extraction tool. Results will be presented as a narrative synthesis with key findings, knowledge gaps, and recommendations. A PRISMA-ScR flow diagram will be included in the final report.

## Introduction

Undernutrition is a major public health issue affecting children worldwide, and can be classified into three main types: stunting (low height for age), wasting (low weight for height), and underweight (low weight for age); a child is diagnosed with any of these ones when its measurement falls below -2 standard deviation of the referenced population (1).

Latin American countries have been able to reduce their child undernutrition prevalence. In 2020, the prevalence was 1.31% for wasting and 2.2% for underweight (2). However, to reduce undernutrition in indigenous children might be a greater challenge. Compared to non-indigenous children, indigenous children have shown a 34% higher prevalence for stunting in Latin america (3); 9% higher risk for undernutrition in Chile (4), and almost 100% higher risk and prevalence for stunting and underweight in Colombia, Peru, and rural communities of Mexico (5–7). The relevance of those findings is not the higher rate itself, the relevance is in the factors causing it. Gatica-Domínguez G. *et al*. (3) expressed that socio-cultural factors might be associated with a higher or lower risk in undernutrition due to ethnicity. Public health workers need to know where they can intervene to improve the nutrition status of these communities, and how the risk factors might differ from the non-indigenous people.

A preliminary search of MEDLINE, the Cochrane Database of Systematic Reviews and JBI Evidence Synthesis was conducted and no current or underway scoping reviews on the topic were identified. There is one systematic review in a similar topic: Batis, C. *et al*.(8) assessed the relationship between stunting and education, wealth, and ethnicity at the ecological level, but their design was not to identify the influence of some risk factors in the prevalence for each of the races. Nowadays, there are more papers, which assess the influence of risk factors between indigenous and non-indigenous children. We need a scoping review to know the gaps, what is currently known, and where a systematic review might provide useful information. That’s why we proposed the followed general objective to research in a scoping review:

*To map the existing literature on the difference in risk factors for undernutrition between indigenous and non-indigenous children younger than 5 years old of Latin America*.

Where the specific objectives will be:

- To identify the main tool measurements used to diagnose undernutrition in indigenous and non-indigenous latin american children younger than 5 years old.
- To explore when indigenous race might be an effect modifier for undernutrition in latin american children younger than 5 years old.
- To explore the possible mechanisms why indigenous latin american children younger than 5 years old might have a higher risk for undernutrition compared to non-indigenous children.

## Review question

*What is known about the difference in risk factors for undernutrition between indigenous and non-indigenous children younger than 5 years old of Latin America and caribbean?*

## Eligibility criteria

### Participants

Indigenous or non-indigenous latin american and caribbean children younger than 5 years old. If the study’s population includes indigenous and non-indigenous children, the results must show stratifications where indigenous infants are not aggregated with non-indigenous children it might be through stratified results or multivariate analyses; also the study must state clearly whether any of its subjects belongs to an indigenous group.

### Concept

Risk factors for child undernutrition, where undernutrition is defined as stunting, wasting, underweight, or a mix of any of them. The study must be designed to find associations between child undernutrition and possible risk factors, that’s why we will exclude any quantitative study that doesn’t have any covariate different to race or ethnicity or uses undernutrition as a risk factor.

### Context

Latin America and Caribbean countries according by the classification of the World bank (9), which includes: Antigua and Barbuda, Argentina, Aruba, Bahamas, Barbados, Belize, Bolivia, Brazil, British Virgin Islands, Cayman Islands, Chile, Colombia, Costa Rica, Cuba, Curaçao, Dominica, Dominican Republic, Ecuador, El Salvador, Grenada, Guatemala, Guyana, Haiti, Honduras, Jamaica, Mexico, Nicaragua, Panama, Paraguay, Peru, Puerto Rico, St. Kitts and Nevis, St. Lucia, St. Martin (French part), St. Vincent and the Grenadines, Suriname, Trinidad and Tobago, Turks and Caicos Islands, Uruguay, Venezuela, RB, U.S. Virgin Islands.

### Types of Sources

This scoping review will consider both experimental and quasi-experimental study designs including randomized controlled trials, non-randomized controlled trials, before and after studies and interrupted time-series studies. In addition, analytical observational studies including prospective and retrospective cohort studies, case-control studies and analytical cross-sectional studies will be considered for inclusion. This review will also consider descriptive observational study designs including case series, individual case reports and descriptive cross-sectional studies for inclusion.

Qualitative studies will also be considered that focus on qualitative data including, but not limited to, designs such as phenomenology, grounded theory, ethnography, qualitative description, action research and feminist research.

In addition, systematic reviews that meet the inclusion criteria will also be considered, depending on the research question.

Text, opinion, and governamental papers will also be considered for inclusion in this scoping review.

## Methods

The proposed scoping review will be conducted in accordance with the JBI methodology for scoping reviews (10).

### Search strategy

The first step of the search strategy will involve a limited search by the review team in PubMed to discover pertinent keywords and search terms that have been utilized in the titles and abstracts of relevant studies within the subject area. The team analyzed the text words in the titles and abstracts of articles identified in the search to develop a full search strategy.

The second step of the search strategy will involve adapting the full search strategy to each database and applying it systematically to several databases including MEDLINE (Pubmed), Embase, Cochrane Library (Ovid), SciELO, EbscoHost (All), LILACS and Epistemonikos. A full search strategy for MEDLINE is presented in Appendix I.

In addition to searching these databases, the review team will also search several trial registries including ClinicalTrials.gov, International Clinical Trials Registry Platform (ICTRP), ISRCTN Registry, and ReBEC (Brazilian Clinical Trials Registry) for relevant studies.

The third step of the search strategy will involve searching for unpublished studies and gray literature using modified search terms via Latin American government repositories, Medxiv and Google Scholar. Finally, the third phase will involve searching for relevant references in the articles identified in step two using Scopus, and conducting manual searches as necessary as mentioned in the Types of Sources.

In addition, we will use the public version of the L.OVE repository of malnutrition which contains systematic reviews related to malnutrition: *https://app.iloveevidence.com/loves/5da3109a69c00e78ea7056d9?question_domain=5b1dcd8ae611de7ae84e8f14&population=5da3104d69c00e78ea7056d8&classification=systematic-review&search=indigenou%2a%20AND%20%20children*

The search strategy will aim to locate both published and unpublished studies. An initial limited search of MEDLINE was undertaken to identify articles on the topic. The text words contained in the titles and abstracts of relevant articles, and the index terms used to describe the articles were used to develop a full search strategy for (see Appendix #1). The search strategy, including all identified keywords and index terms, will be adapted for each included database and/or information source. The reference list of all included sources of evidence will be screened for additional studies.

Studies published in any language will be included. Studies published since 2000 will be included.

### Study/Source of Evidence selection

Following the search, all identified citations will be collated and uploaded into *Zotero version No. 6* and duplicates removed. Titles and abstracts will then be screened by two or more independent reviewers for assessment against the inclusion criteria for the review using Rayyan. Any discrepancies that arise will be resolved through discussion or by seeking the opinion of a third reviewer. All studies included in the title/abstract screening will be obtained in full-text format. The full text of included studies will be assessed in detail against the inclusion criteria by two or more independent reviewers. Reasons for exclusion of sources of evidence at full text that do not meet the inclusion criteria will be recorded and reported in the scoping review. Any disagreements that arise between the reviewers at each stage of the selection process will be resolved through discussion, or with an additional reviewer/s. The results of the search and the study inclusion process will be reported in full in the final scoping review and presented in a Preferred Reporting Items for Systematic Reviews and Meta-analyses extension for scoping review (PRISMA-ScR) flow diagram (11).

### Data Extraction

Data will be extracted from papers included in the scoping review by two or more independent reviewers using a data extraction tool developed by the reviewers. The data extracted will include specific details about the participants, concept, context, study methods, study outcomes, and key findings relevant to the review question/s.

A draft extraction form is provided (see Appendix II). The draft data extraction tool will be modified and revised as necessary during the process of extracting data from each included evidence source. Modifications will be detailed in the scoping review. Any disagreements that arise between the reviewers will be resolved through discussion, or with an additional reviewer/s. If appropriate, authors of papers will be contacted to request missing or additional data, where required.

### Data Analysis and Presentation

Results will be presented as a narrative synthesis, along with key findings, identified knowledge gaps, and recommendations. Depending on the key findings, the outcomes will be presented in charted or tabular form outlining the characteristics of included sources of evidence and displaying the frequency of their outcomes.

## Data Availability

All data produced in the present study are available upon reasonable request to the authors

## Acknowledgements

*None*

## Appendices

### Appendix I: Search strategy

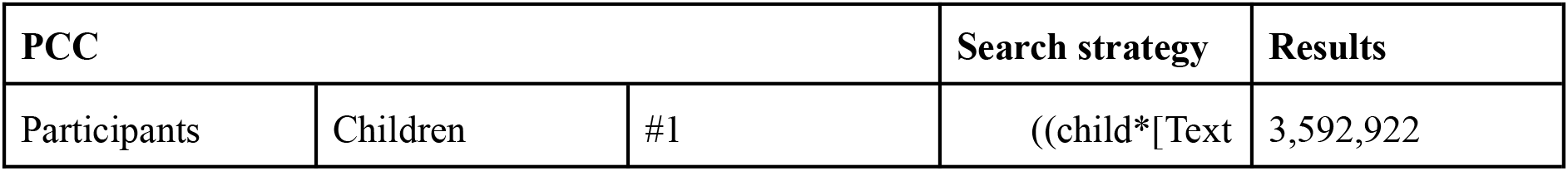
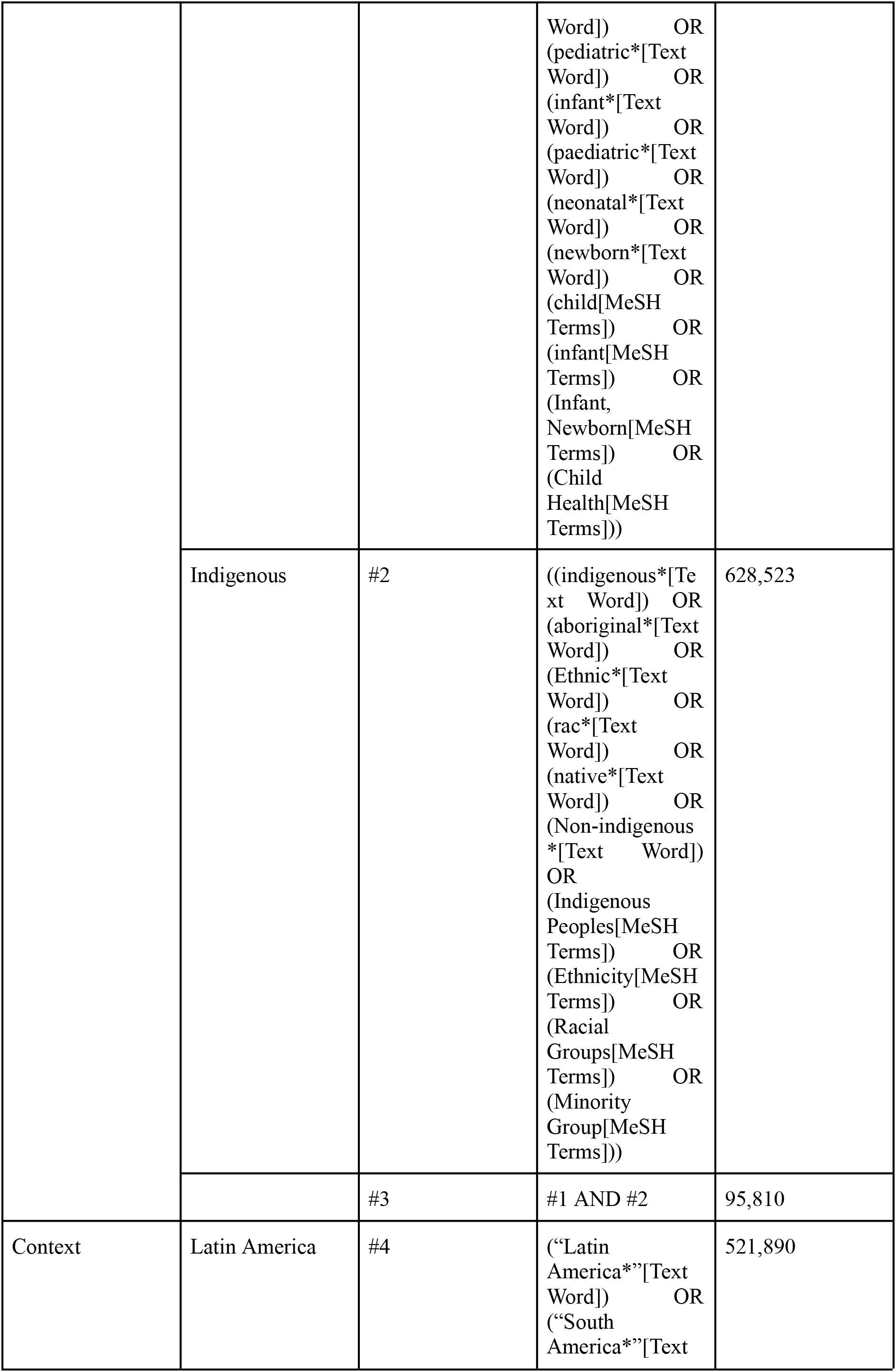
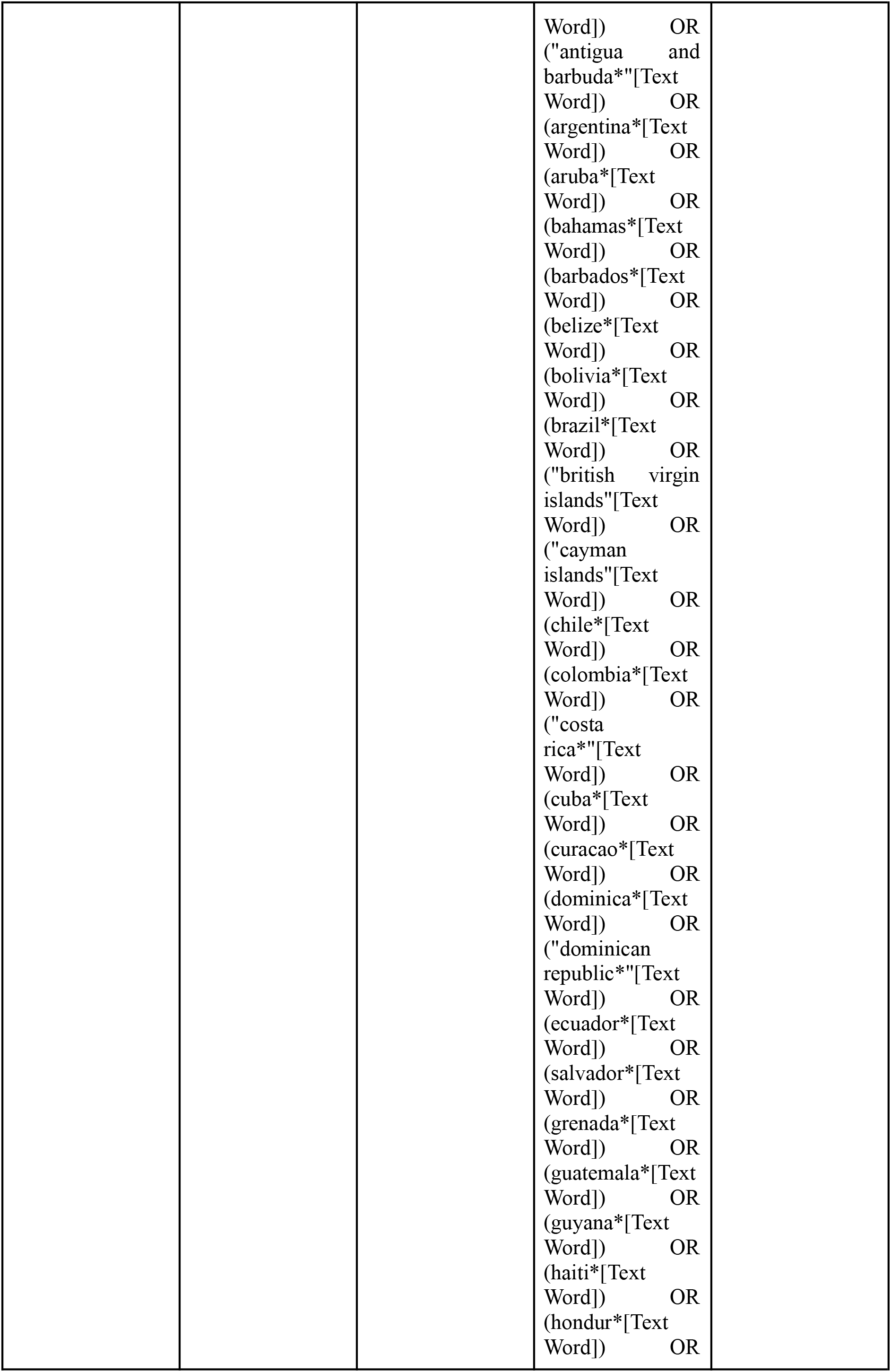
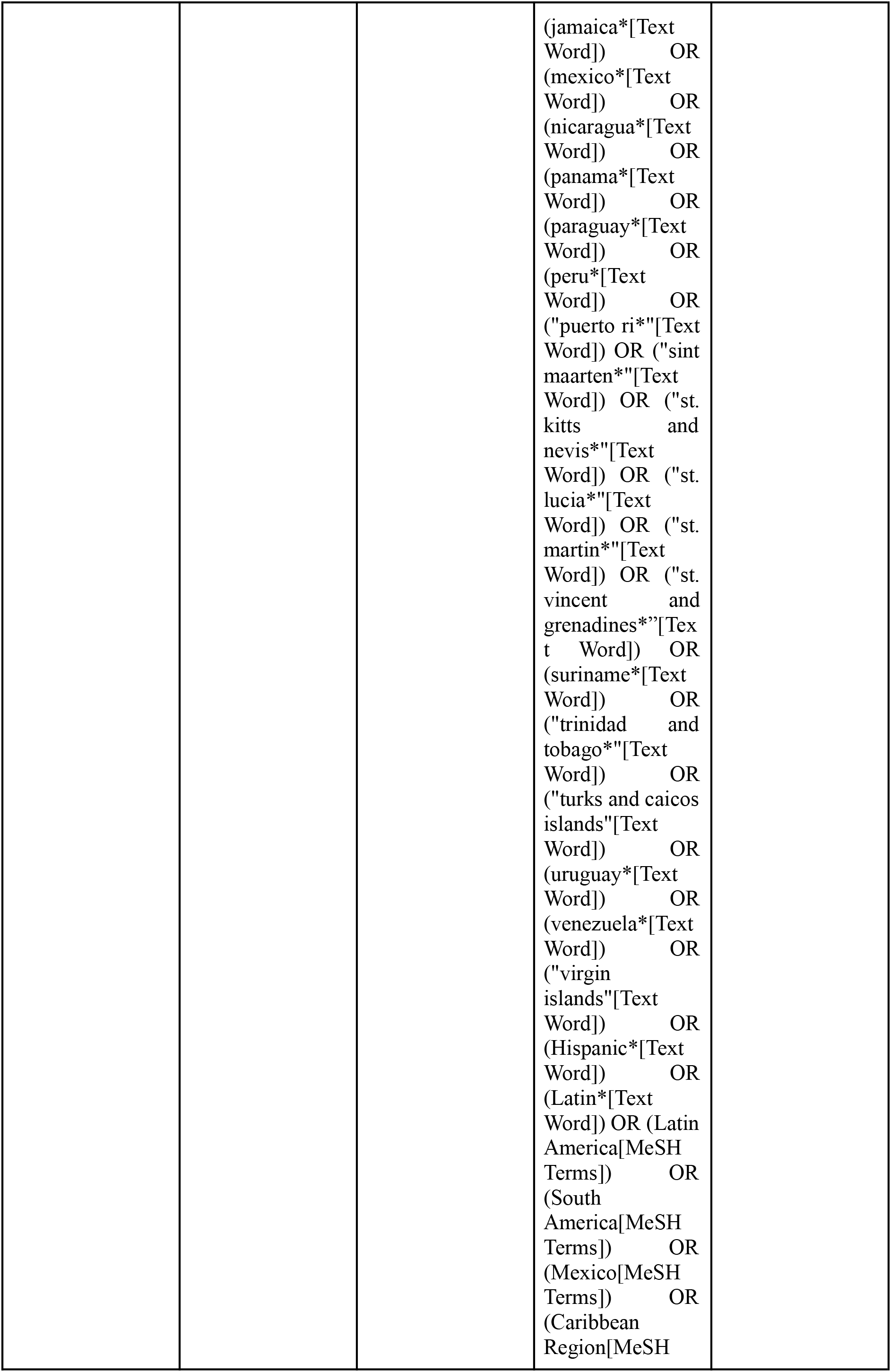
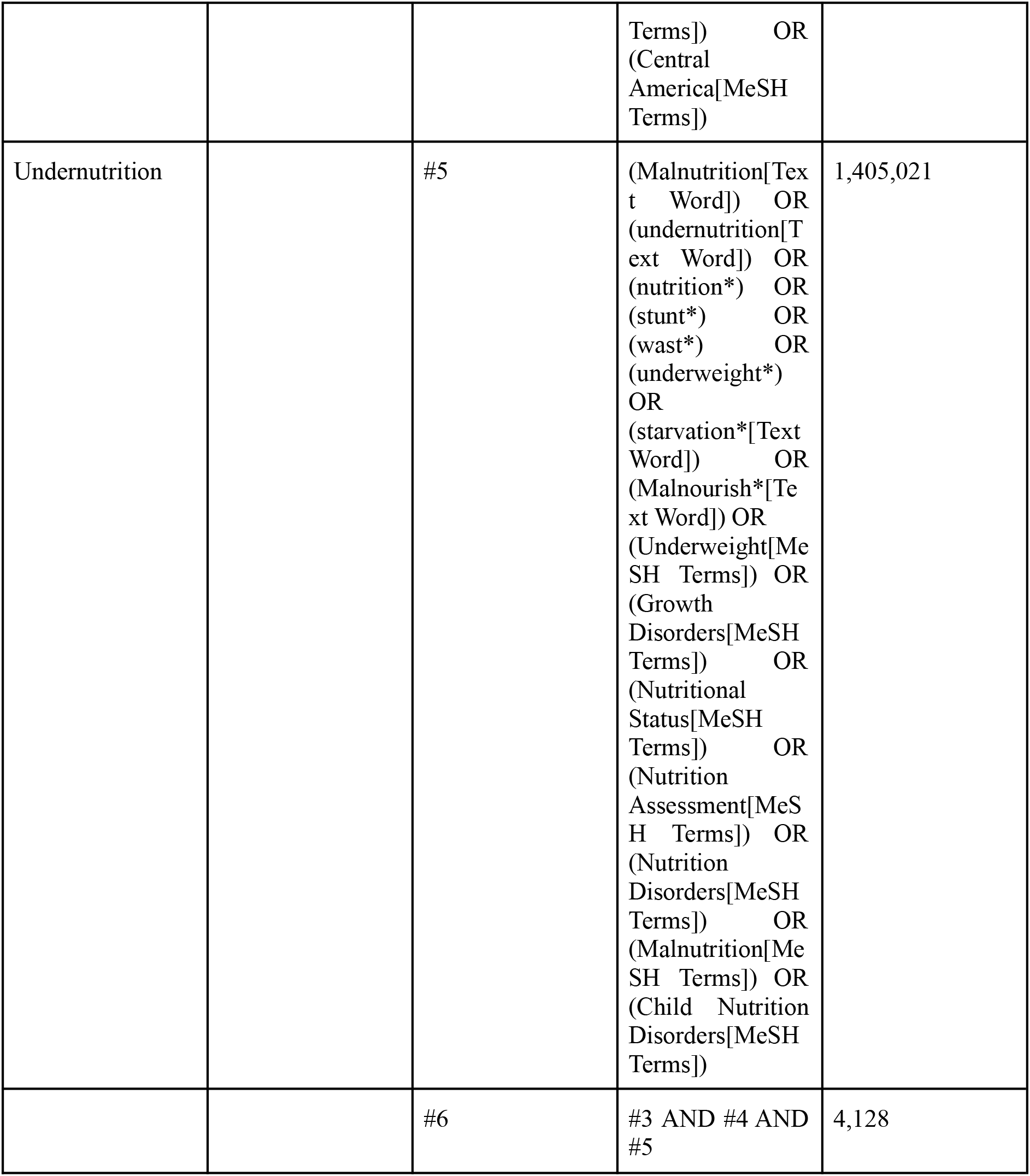

### Appendix II: Data extraction instrument

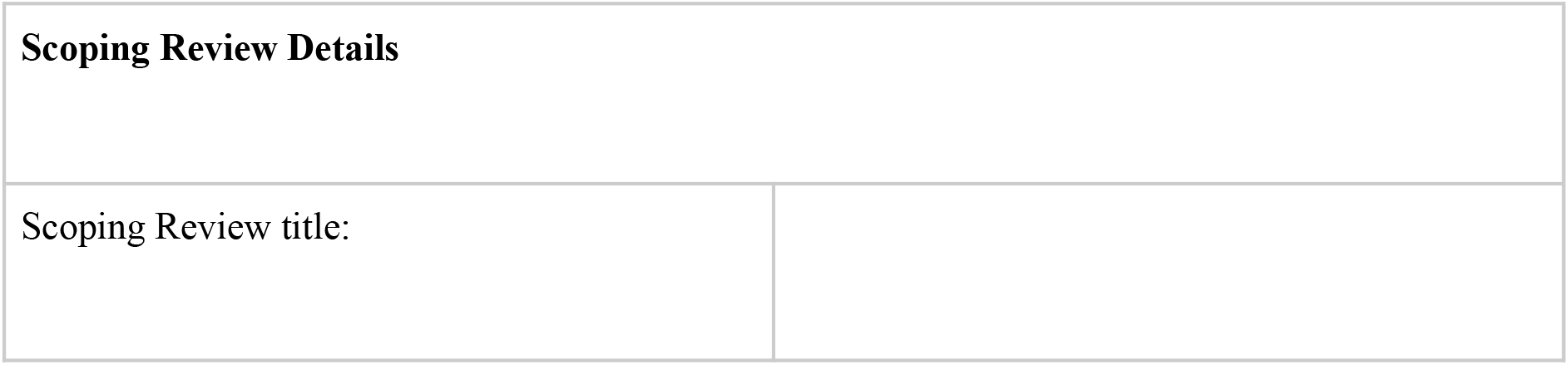
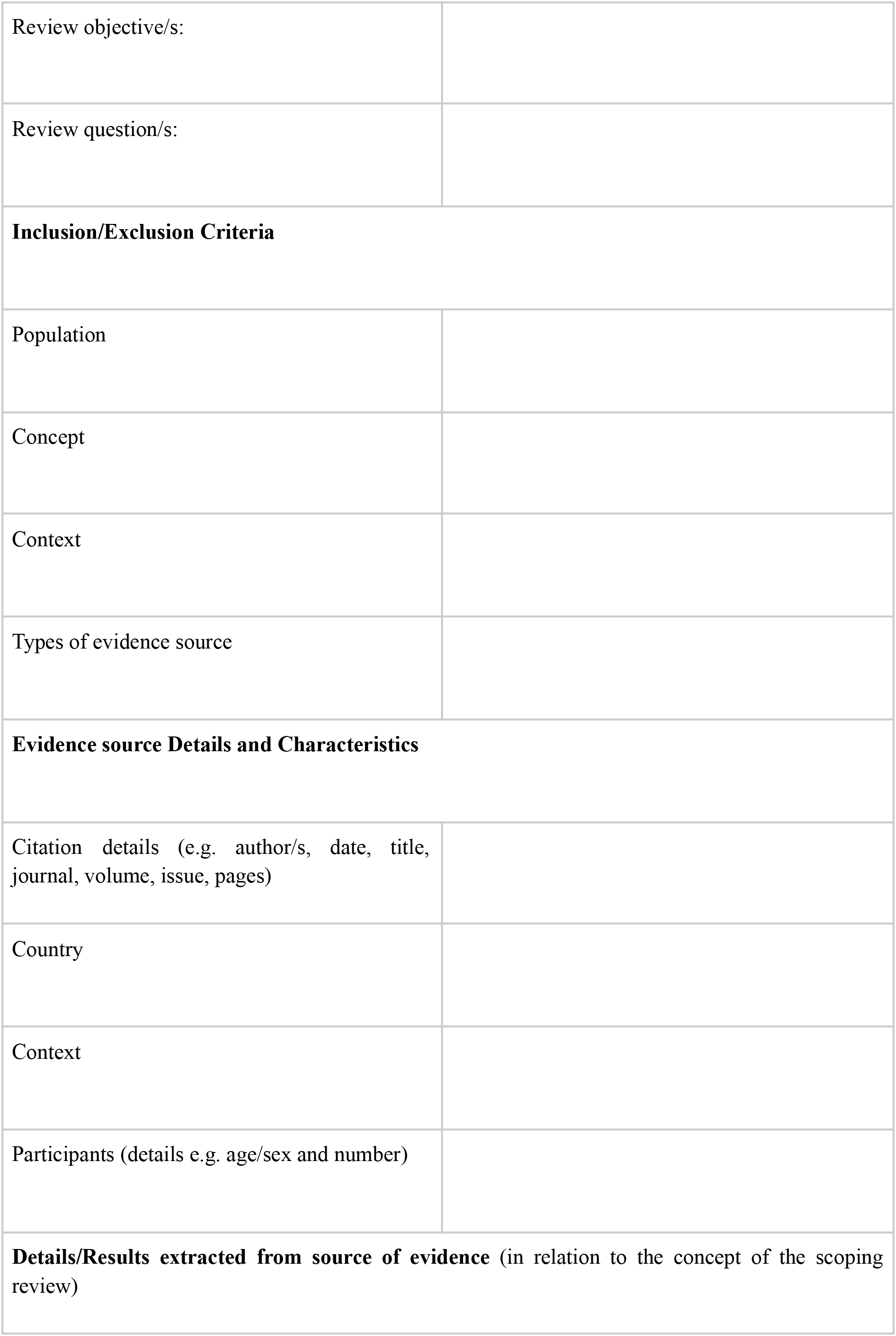

